# Artificial intelligence–enabled electrocardiogram for mortality and cardiovascular risk estimation: An actionable, explainable and biologically plausible platform

**DOI:** 10.1101/2024.01.13.24301267

**Authors:** Arunashis Sau, Libor Pastika, Ewa Sieliwonczyk, Konstantinos Patlatzoglou, Antonio H. Ribeiro, Kathryn A. McGurk, Boroumand Zeidaabadi, Henry Zhang, Krzysztof Macierzanka, Danilo Mandic, Ester Sabino, Luana Giatti, Sandhi M Barreto, Lidyane do Valle Camelo, Ioanna Tzoulaki, Declan P. O’Regan, Nicholas S. Peters, James S. Ware, Antonio Luiz P. Ribeiro, Daniel B. Kramer, Jonathan W. Waks, Fu Siong Ng

## Abstract

**Background and Aims:** Artificial intelligence-enhanced electrocardiograms (AI-ECG) can be used to predict risk of future disease and mortality but has not yet been adopted into clinical practice. Existing model predictions lack actionability at an individual patient level, explainability and biological plausibility. We sought to address these limitations of previous AI-ECG approaches by developing the AI-ECG risk estimator (AIRE) platform.

**Methods and Results:** The AIRE platform was developed in a secondary care dataset of 1,163,401 ECGs from 189,539 patients, using deep learning with a discrete-time survival model to create a subject-specific survival curve using a single ECG. Therefore, AIRE predicts not only risk of mortality, but *time-to-mortality*. AIRE was validated in five diverse, transnational cohorts from the USA, Brazil and the UK, including volunteers, primary care and secondary care subjects. AIRE accurately predicts risk of all-cause mortality (C-index 0.775 (0.773-0.776)), cardiovascular (CV) death 0.832 (0.831-0.834), non-CV death (0.749 (0.747-0.751)), future ventricular arrhythmia (0.760 (0.756-0.763)), future atherosclerotic cardiovascular disease (0.696 (0.694-0.698)) and future heart failure (0.787 (0.785-0.889))). Through phenome- and genome-wide association studies, we identified candidate biological pathways for the prediction of increased risk, including changes in cardiac structure and function, and genes associated with cardiac structure, biological aging and metabolic syndrome.

**Conclusion:** AIRE is an actionable, explainable and biologically plausible AI-ECG risk estimation platform that has the potential for use worldwide across a wide range of clinical contexts for short- and long-term risk estimation.

**Graphical Abstract:** 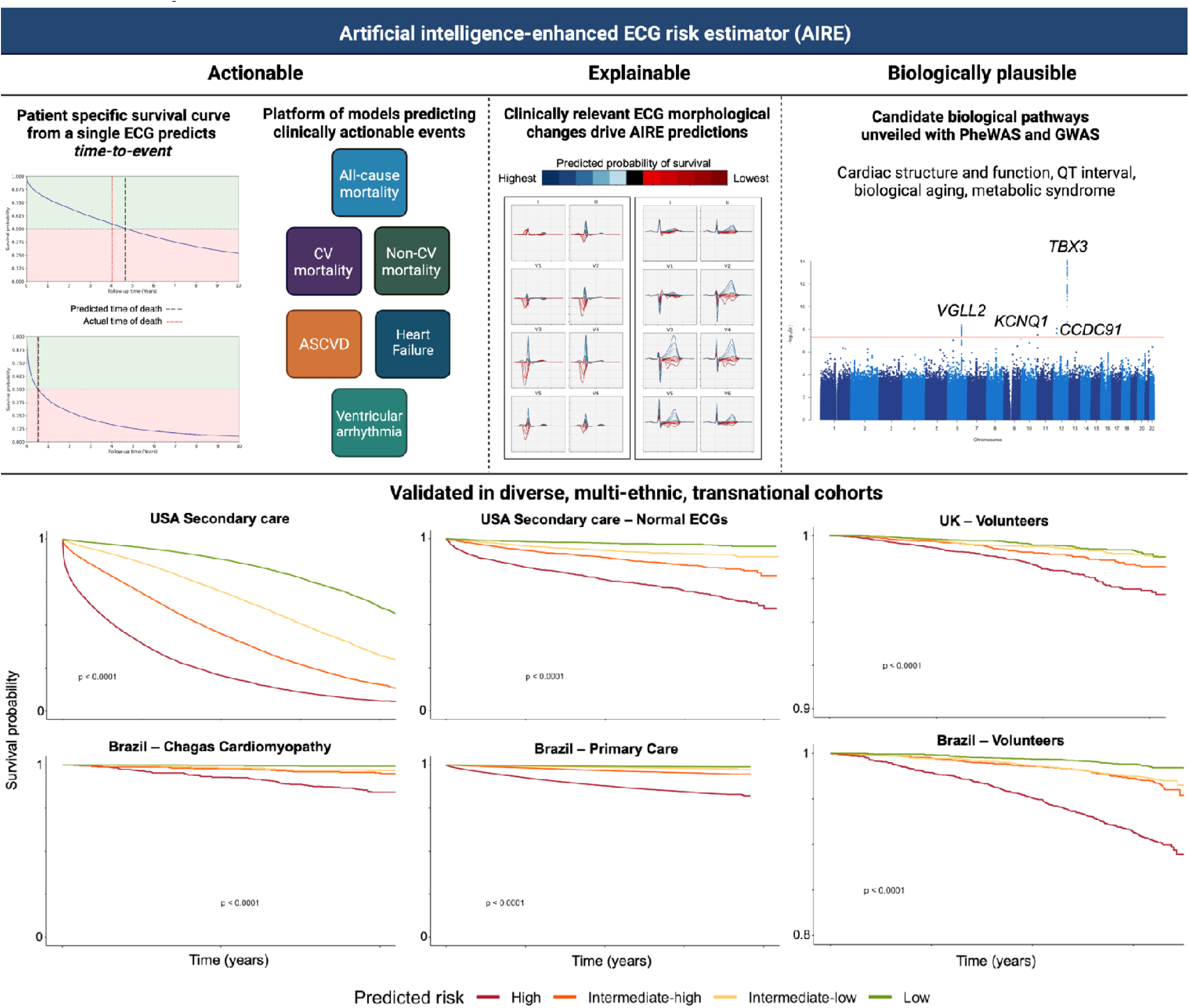

## Introduction

The electrocardiogram (ECG) has been a fundamental tool in clinical medicine for over a century. With the recent advent of artificial intelligence (AI), and in particular deep learning, the potential applications of the ECG have significantly expanded, including both diagnostic and predictive capabilities (1–3). A major advantage of the deep learning approach is the ability to extract features relevant to the specific task, without anchoring on prior human beliefs. Recent studies have demonstrated the remarkable predictive capabilities of AI-ECG models, not only in terms of predicting mortality, but also future cardiac diseases (4–7). Novel AI-ECG biomarkers, such as AI-ECG predicted age and BMI (8, 9), have also been shown to be capable of capturing information on future risk.

However, although many of these existing risk prediction models have good performance metrics, they have not been integrated into routine clinical care. Existing mortality prediction models are limited by prediction of survival at a small number of set time points, rather than providing an individualised survival prediction over time. Just as the Cox proportional hazards model is superior to logistic regression when time-to-event information is available (10), survival-based deep learning may outperform classification-based approaches.

Another key limitation of existing models is the lack of information for clinicians on specific actionable pathways. A high-risk prediction is unhelpful to a clinician if there is no accompanying information on how to affect the survival trajectory of their patient. To make AI-ECG predictions more actionable, it is essential to consider not only time-to-event predictions, but also specific predictions for diseases with established preventive and disease modifying treatments.

Furthermore, the adoption of AI into clinical practice is significantly limited by concerns regarding explainability and biological plausibility. Just as knowledge of the mechanisms of action of drugs are important for physicians to have confidence in their application, biological plausibility of AI predictions ensures their credibility and acceptance.

To address these limitations of existing risk prediction models, we aimed to develop and perform transnational validation on an AI-ECG risk prediction platform that is not only accurate, but also actionable, explainable, and biologically plausible.

## Methods

In this study we first developed the AI-ECG risk estimation (**AIRE**) model for prediction of all-cause mortality. We subsequently developed seven additional submodels. The eight models together are referred to as the **AIRE platform**. A model development and validation flow chart is shown in **Figure 1**.

**Figure 1.**
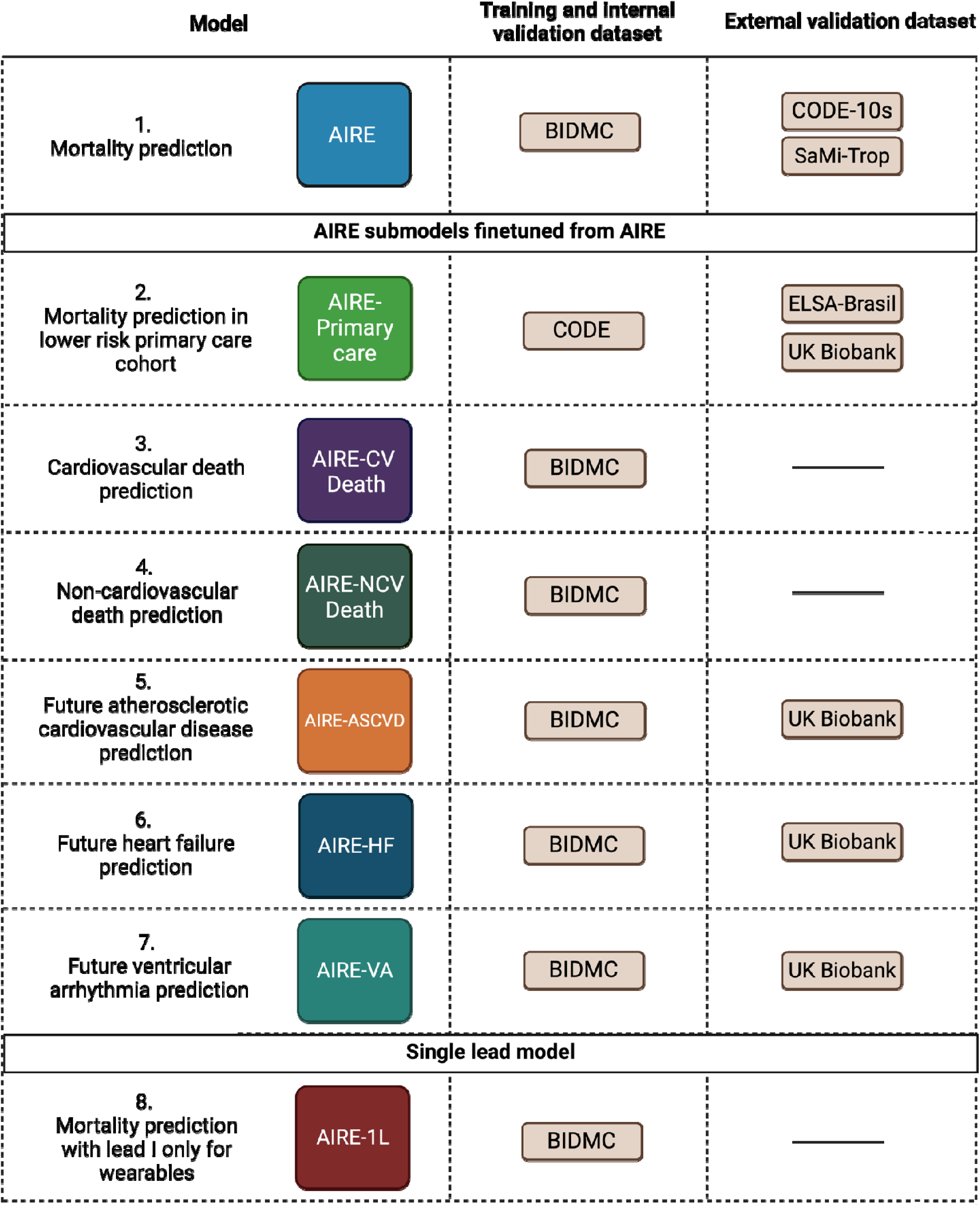
Schematic depicting all eight models in the **AIRE platform**, training datasets and validation datasets. **AIRE** and **AIRE-primary care** were trained for all-cause mortality, the remaining models were trained for the outcomes they are named after. CODE-10s denotes the subset of CODE with ECGs of 10s duration. CV: cardiovascular, NCV: non-cardiovascular, ASCVD: atherosclerotic cardiovascular disease, HF: heart failure, VA: ventricular arrhythmia. BIDMC: Beth Israel Deaconess Medical Center, CODE: Clinical Outcomes in Digital Electrocardiography, SaMi-TROP: São Paulo-Minas Gerais Tropical Medicine Research Center, ELSA-Brasil: The Brazilian Longitudinal Study of Adult Health.

### Ethical approvals

This study complies with all relevant ethical regulations, further details are provided in the **Supplementary Methods**.

### Cohorts

We studied five cohorts, briefly the Beth Israel Deaconess Medical Center (BIDMC) cohort is a secondary care dataset comprised of routinely collected data from, Boston, USA. The São Paulo-Minas Gerais Tropical Medicine Research Center (SaMi-Trop) is a cohort of patients with chronic Chagas cardiomyopathy (11). The Clinical Outcomes in Digital Electrocardiography (CODE) cohort is a Brazilian database of ECGs recorded in primary care (8) containing ECGs of both 10s and 7s duration. The subset of this dataset with only 10s ECGs is referred to as CODE-10s. The Longitudinal Study of Adult Health (ELSA-Brasil) cohort consists of Brazilian public servants (12). The UK Biobank (UKB) is longitudinal study of volunteers (13). Further details are provided in the **Supplementary Methods**.

### ECG pre-processing

12-lead ECGs were pre-processed with a bandpass filter 0.5 to 100Hz, a notch filter at 60Hz and re-sampling to 400Hz. Zero padding was added to make the input shape a power of 2. This resulted in 4096 samples for each lead for a 10s recording (4000 samples + 48 zeros at the start and end), that was used as input to the neural network model. As leads III, aVL, aVR, aVF are linear combinations of leads I and II, these leads were not used for model development or evaluation. Therefore, the final input shape of a single ECG was 4096 x 8.

### AI-ECG risk estimation platform development

We developed the **AIRE platform** using the BIDMC cohort as the derivation dataset. The data was split at a ratio of 50/10/40% for training, validation and internal test, respectively. For mortality end-points, ECGs without paired life status at 30 days were excluded. Data was split by patient ID stratified by presence of ECGs with paired 5-year life status. To prevent data leakage, a single subject could have ECGs assigned to only one of training/validation/testing datasets. We used a previously described convolutional neural network architecture based on residual blocks (14) and adapted the final layer to accommodate a discrete-time survival model (15). The discrete-time survival approach allows the model to account for both time to outcome (mortality) and censorship (i.e., loss to follow up). Unlike other models trained to predict mortality at one, or a small number of time points, our model predicts outcomes at numerous timepoints up to 10 years and accounts for right censored data during model training.

The lead I model, **AIRE-1L**, was developed using the same methodology above, using the same BIDMC data split but using just lead I as the model input. The Transparent Reporting of a Multivariable Prediction Model for Individual Prognosis or Diagnosis Checklist for Prediction Model Development and Validation was followed (16).

### Model fine-tuning for primary care population

Using the CODE dataset, we finetuned the model to be more representative of a primary care population (**AIRE-primary care**) with a much lower risk of adverse events. We used 75% of the CODE dataset for finetuning, with 5 % as a validation set. The final 20% was used for internal validation of **AIRE-primary care**.

### Model fine-tuning for other endpoints

We also developed five other subsequent models by fine-tuning the AIRE model separately for cardiovascular (CV) death, non-CV death, ventricular arrhythmia (VA), atherosclerotic cardiovascular disease (ASCVD) and heart failure (HF) in the BIDMC dataset. These models were named **AIRE-CV death**, **AIRE-NCV death**, **AIRE-VA**, **AIRE-ASCVD** and **AIRE-HF** respectively. The same splits were used as for training the original model. Fine-tuning was performed by loading the previous model and training using a low learning rate without freezing any layers. Internal validation and external validation datasets are shown in **Figure 1**.

For non-mortality endpoints (ASCVD, VA, HF), subjects were coded as having prevalent disease, future disease or neither at the time of the ECG. Although the goal of these models was to predict future events, we hypothesised that information on prevalent disease would be helpful for model training. In order to include prevalent disease in the discrete-time survival model, we encoded prevalent disease at the first timepoint in the discrete-time survival labels. When evaluating model performance, subjects with prevalent disease were excluded.

### Comparison of AIRE with other models

We compared **AIRE and AIRE-primary care** to the previously described AI-ECG predicted age (8). AI-ECG predicted age values are publicly available for CODE15 and SaMi-Trop. The remaining 85% of CODE was used to develop the AI-ECG age model, and therefore was not used. For ELSA-Brasil, the model weights were downloaded (17) and used to derive AI-ECG predicted age values.

We also compared **AIRE-primary care** and **AIRE-ASCVD** to the recently described SEER (Stanford Estimator of ECG Risk), which had a similar goal of predicting cardiovascular mortality and ASCVD (7). The model code and weights were downloaded (18) and performance evaluated in the UKB, as this was a dataset external to **AIRE-primary care, AIRE-ASCVD** and SEER, with cause of death and ASCVD event data available.

### Survival and statistical analyses

In the test set, we generated predictions of all ECGs for the primary analyses of all-cause mortality. Sensitivity analyses including a single random ECG per subject were also performed. Model performance was reported using the C-index, and time-dependent AUROC. For analyses requiring a single predictor value (C-index and AUROC), the probability of survival at 5 years was used. Risk quartiles were defined (low, intermediate-low, intermediate-high, high) based on values in the validation set. Given the diverse populations and event rates evaluated, risk quartiles were redefined in each dataset. In each case, where categorical risk levels were required, 5% of the dataset was used to define the quartiles and evaluation was performed in the remaining 95%. Kaplan-Meier curves comparing the risk quartiles were plotted and statistical significance assessed using the log rank test.

Cox models were fit using the test dataset comparing demographics, clinical variables, imaging parameters and **AIRE platform** predictions. For the Cox models incorporating **AIRE platform** predictions, all model outputs (i.e., predicted probabilities of death at each timepoint) were used as inputs, as well as age, sex, heart rate, PR interval, QRS duration and QTc interval. These models are designated AIRE-Cox for the AIRE model and AIRE-CV Death-Cox, AIRE-ASCVD-Cox, AIRE-VA-Cox, AIRE-HF-Cox for the other models. Complete case analysis was used, therefore no variables were imputed. Recent work suggests virtually all real-world clinical datasets will violate the proportional hazards assumptions if sufficiently powered and that statistical tests for the proportional hazards assumption may be unnecessary (19). In line with these recommendations, the proportional hazards assumption was not evaluated and the hazard ratio from our Cox models should be interpreted as a weighted average of the true hazard ratios over the follow-up period. Nested Cox models were compared with the Likelihood Ratio test, while non-nested Cox models were compared with the Partial Likelihood Ratio test (20). Statistical analyses were performed with R 4.2.0 statistical package (R Core Team, Vienna, Austria) or Python (version 3.9).

### Diagnostic and imaging data

ICD-9 and ICD-10 codes were used to define presence/absence of disease in the BIDMC and UKB cohorts. Cardiovascular death in the BIDMC cohort was defined as mortality occurring within 30 days of a diagnostic code for acute myocardial infarction, ischaemic stroke, intracranial haemorrhage, sudden cardiac death, or heart failure as previously described (7, 21). In the UKB, cause of death was ascertained based on the ICD10 code stated as the primary cause of death. Diagnostic codes were not available in the SaMi-Trop, CODE and ELSA-Brasil datasets. Echocardiograms within 60 days of an ECG were linked and used for analyses incorporating echocardiographic parameters. The pooled cohort equation was calculated using the PooledCohort R package. Medication usage was not available in the BIDMC cohort, therefore ICD9 and ICD10 codes consistent with a diagnosis of hypertension were used to code for antihypertensive medication use for calculation of the pooled cohort equation. Blood results and blood pressure (BP) readings taken within 180 days of the ECG were averaged. Sensitivity analyses were performed using 90 days and 30 days results.

We investigated the performance of AIRE in the high risk disease groups of severe aortic stenosis and primary pulmonary hypertension. Severe aortic stenosis was defined based on the reported overall severity on echocardiography reports (i.e. a subjective overall assessment by the clinician undertaking the echocardiogram). Primary pulmonary hypertension was defined using ICD9 and ICD10 codes. No fine-tuning was performed when evaluating the performance of **AIRE** in the severe aortic stenosis and primary pulmonary hypertension disease groups.

### Normal ECG definition

We evaluated the performance of AIRE in clinician reported normal ECGs. In the BIDMC dataset a subset of ECGs had Cardiologist reports. Normal ECGs in BIDMC were determined by searching for ‘normal ecg’ in the free text reports, a whole word match was required in order to exclude ‘abnormal ecg’. ECGs with the phrase ‘otherwise’ were also excluded from the normal definition. Additionally we filtered by heart rate (60-100 bpm), PR interval (less than 200ms), QRS duration (less than 120ms) and QTc interval (less than 470ms). Normal ECGs in ELSA-Brasil and CODE were defined as previously described (17).

### Explainability

In order to understand the ECG morphologies associated with predicted survival we used two approaches. Median beats were extracted using the BRAVEHEART ECG analysis software as previously described (22).

First, we trained a variational autoencoder (VAE) as previously described (23) using median ECG beats. Further details in **Supplementary Methods.** In preliminary analyses, models based on only the VAE latent features were found to be inferior to the supervised deep learning approach described above, therefore the VAE was used for explainability only, and not used for **AIRE** model training or any of the prediction models described in this manuscript. VAE latent features were input into a linear regression with predicted survival as the output. The top 3 most important features as assessed by the t-value were visualised by latent traversal (23).

Second, using the median beats we calculated the average waveform from the 10,000 ECGs with the lowest and highest **AIRE** predicted mortality. The mean and standard deviation of these waveforms was then plotted.

### PheWAS

To better understand the biology underlying **AIRE** and to explore the detailed phenogroup associations, we performed phenome-wide association studies (PheWAS). We performed PheWAS analysis in the UKB, which contains data from over 3000 phenotypes derived from patient measurements, surveys, and investigations. Univariate correlation was performed to investigate the association between ECG phenogroup and phenotypes, adjusted for age, sex and age^2^. We additionally investigated the association of predicted survival with continuous echo traits in the BIDMC dataset. Left ventricular trabeculation was calculated as previously described (24). Deep learning-derived brain age was calculated as previously described (25).

### GWAS

To identify genetic associations with the ECG phenogroups, we performed a genome-wide association study (GWAS) in the UKB. As the predicted survival trait was skewed, the data were normalized by rank-based inverse normal transform prior to the analysis. The GWAS analysis was adjusted for the following covariates: age at imaging visit, sex, height, body mass index (BMI), imaging assessment centre and the first 10 genetic principal components.

Further methods are described in the **Supplementary Methods**.

## Results

### AIRE accurately predicts mortality across diverse timepoints

In the BIDMC cohort, 1,163,401 ECGs were available from 189,539 subjects. Mean follow-up period was 5.46±5.81 years on a per ECG basis, 3.41 (4.08) years taking a random ECG per subject. 34,938 (18.4%) subjects died during follow-up (**Table 1**).

**Table 1.** Dataset demographics. Data at the timepoint of a randomly selected ECG per subject is shown for the BIDMC and CODE datasets. Categorical variables n (%), continuous variables mean (SD)

|  | BIDMC | SaMi-TROP | CODE | ELSA-Brasil | UK Biobank |
| --- | --- | --- | --- | --- | --- |
| N subjects | 189539 | 1631 | 1558421 | 13739 | 42386 |
| Age | 57.68 (18.69) | 59.40 (12.79) | 51.66 (17.59) | 52.26 (9.10) | 64.14 (7.75) |
| Follow up (years) | 3.41 (4.08) | 2.08 (0.39) | 3.68 (1.87) | 9.35 (1.28) | 3.73 (1.57) |
| Sex (M) | 90792 (47.9) | 534 (32.7) | 627042 (40.2) | 7483 (54.5) | 20538 (48.5) |
| Hypertension | 74409 (39.3) | - | 492640 (31.6) | 4924 (35.9) | 6256 (14.8) |
| Previous MI | 11788 (6.2) | - | 11604 (0.7) | 251 (1.8) | 633 (1.5) |
| Smoker | 23343 (12.3) | - | 108815 (7.0) | 1801 (13.1) | 1111 (2.6) |
| Diabetes Mellitus | 33748 (17.8) | - | 101470 (6.5) | 2728 (19.9) | 1334 (3.1) |
| Hyperlipidaemia | 67087 (35.4) | - | 60590 (3.9) | 6811 (49.6) | 2985 (7.0) |
| Mortality | 34938 (18.4) | 104 (6.4) | 52127 (3.3) | 599 (4.4) | 526 (1.2) |

**Table 2.** Mortality prediction results summary table. Model performance as assessed by C-index (95% CI) is shown **AIRE**: Artificial-intelligence enhanced ECG risk estimator

|  | **BIDMC Test set** | **SaMi-Trop** | **CODE-10s** | **CODE Test set** | **UK Biobank** | **ELSA-Brasil** |
| --- | --- | --- | --- | --- | --- | --- |
| **N ECGs** | 434,629 | 1631 | 958,954 | 464,429 | 42,386 | 13,739 |
| **N Subjects** | 64,105 | 1631 | 645,372 | 311,684 | 42,386 | 13,739 |
| **Internal/external validation** | Internal | External | External | Internal | External | External |
| **AIRE** | 0.775 (0.773-0.776) | 0.773 (0.733-0.813) | 0.762 (0.759-0.765) | - | - | - |
| **AIRE-primary care** | - | - | - | 0.802 (0.799-0.805) | 0.638 (0.608-0.668) | 0.713 (0.691-0.735) |

**AIRE** produces subject-specific survival curves from only a single ECG and can predict *time-to-death.* **Figure 2A** and **2B** demonstrate two representative subject-specific survival curves in patients who died during follow up and two curves from subjects who survived through the follow up period, each generated by **AIRE** from a single ECG. **Figure 2C** demonstrates the evolution of AIRE-predicted survival based on multiple ECGs performed over several years of follow up. ECGs nearer to the subject’s time of death show falling survival probabilities, particularly shortly before the subject’s death.

**Figure 2.**
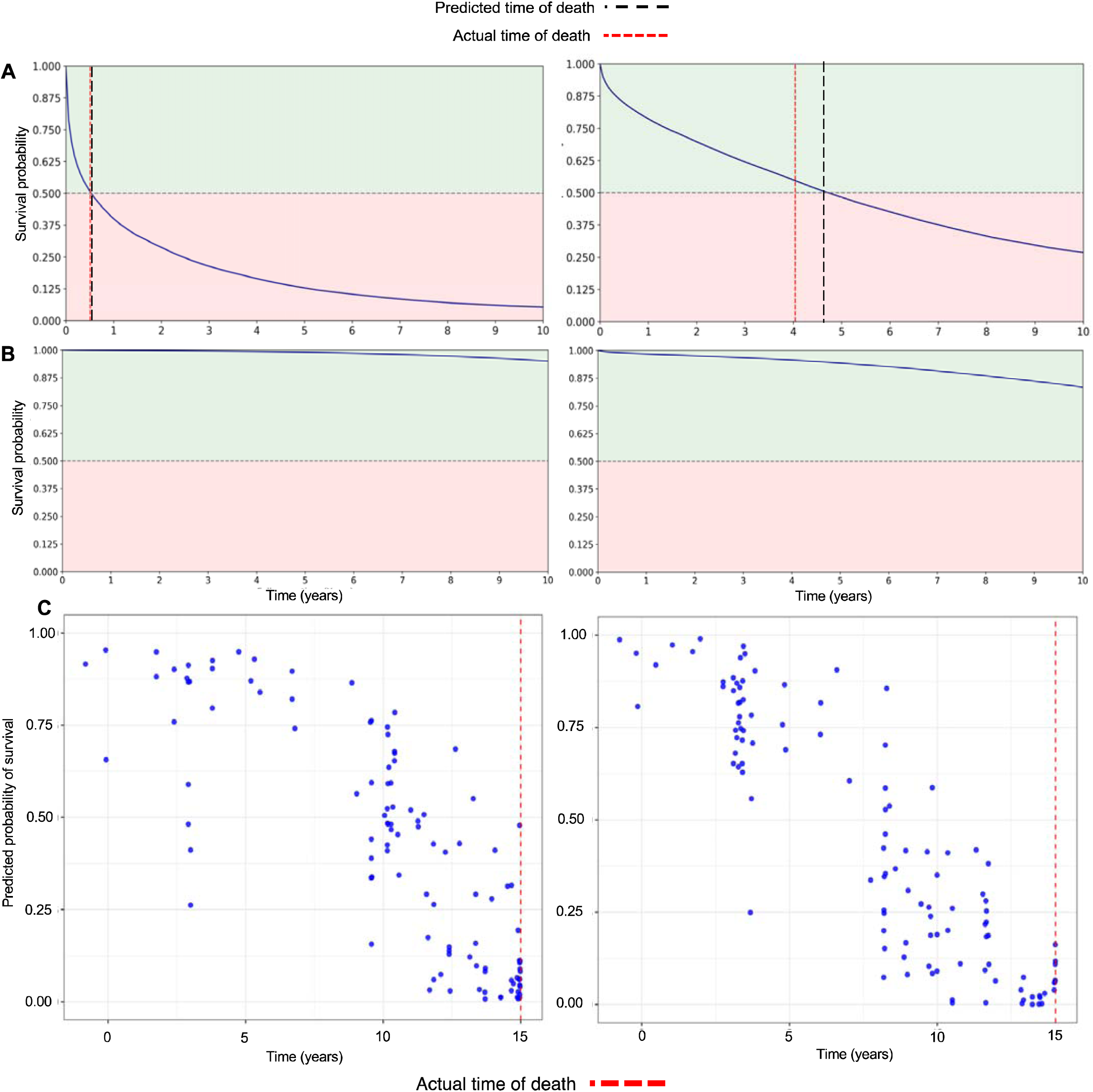
Example subject-specific survival predictions. **AIRE** outputs subject-specific survival curves. Two examples are shown for subjects who died during follow up (A), and two for subjects who survived through the follow up period (B). Dashed red lines indicate the date of death and dashed black lines indicate **AIRE**-predicted date of death. (C) Examples of subjects with many ECGs during the study period, each blue dot is a survival prediction from a single ECG. **AIRE**-predicted survival trends down over time and predicted probability of survival is particularly low prior to actual time of death (red dashed line).

In the hold out test set, **AIRE** predicted all-cause mortality with a concordance-index of 0.775 (0.773-0.776), further results are reported in the **Supplementary Results**. Using the predicted probability of survival at 5 years in the validation set, quartiles of risk (low, intermediate-low, intermediate-high and high) were determined. **Figure 3A** shows the marked separation of survival curves of these quartiles in the test set. **Table S1** shows age and sex adjusted hazard ratios for high risk vs low risk quartile for all cohorts.

**Figure 3.**
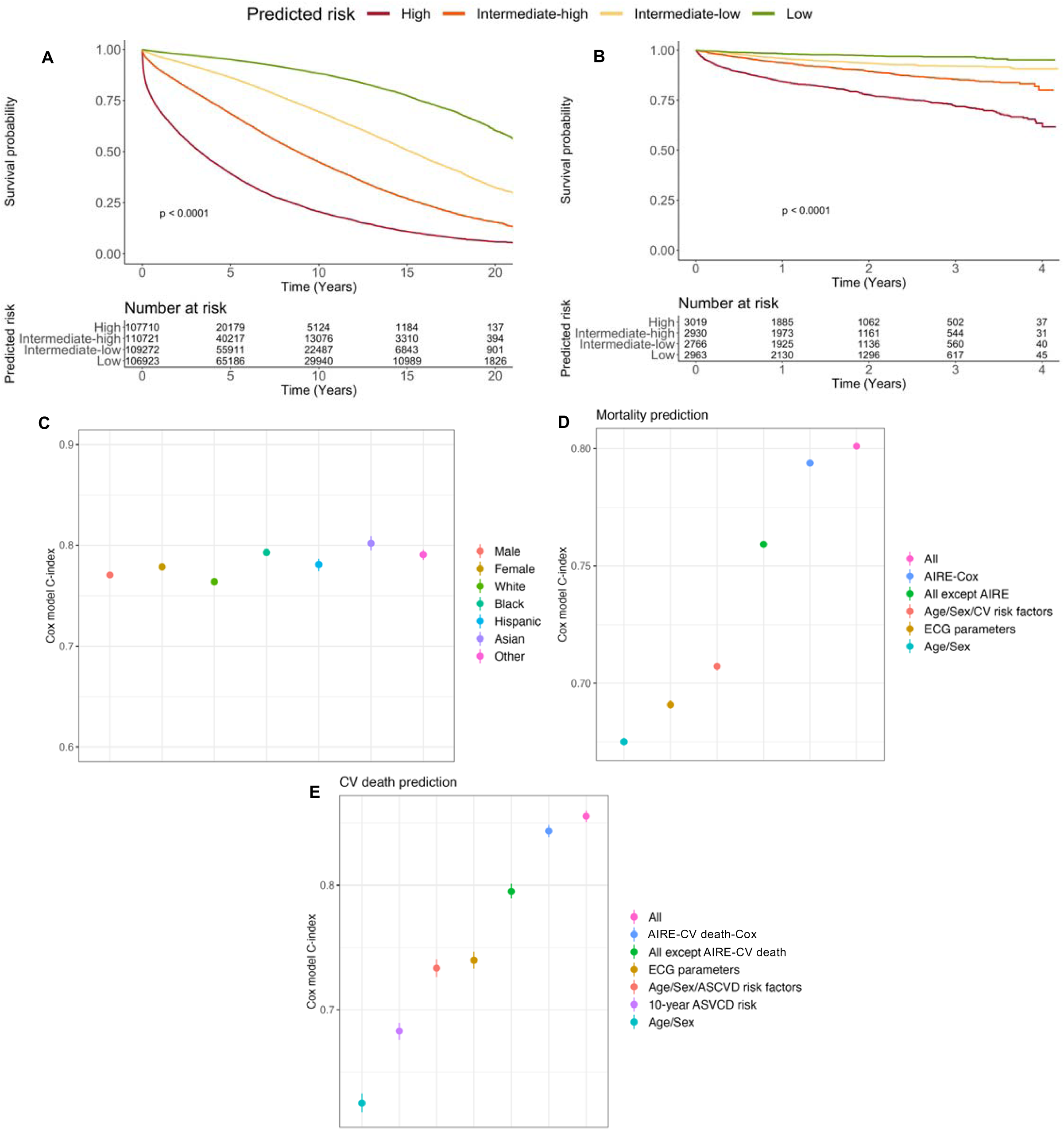
Mortality prediction performance – BIDMC Test set. Kaplan-Meier curves of **AIRE**-predicted all-cause mortality by risk quartile in the whole BIDMC test set (A) and a subset of normal ECGs (B). (C) Comparison of **AIRE** performance across sex and major ethnic groups, **AIRE** performs well across all demographic groups. Using Cox models, **AIRE** was compared with existing risk factors and ECG parameters. In these Cox models, age, sex and ECG parameters were incorporated with **AIRE** to create **AIRE-Cox**. In all comparisons, **AIRE-Cox** had a significantly higher C-index than all comparators for both all-cause mortality (D), and cardiovascular mortality (E). ECG parameters: heart rate, PR interval, QRS duration, QTc interval, CV risk factors: diabetes mellitus, hypertension, smoking history, hyperlipidaemia, ethnicity. ASCVD risk factors: systolic blood pressure, total cholesterol, HDL cholesterol, hypertension, smoking history, diabetes mellitus, ethnicity. 10-year ASCVD risk assessed using the pooled cohort equation.

A subset of ECGs had cardiologist interpretations available. These were more recent ECGs with shorter follow up durations compared to the whole cohort. When considering only ECGs labelled as normal by cardiologists, there remained a significant difference in mortality between high-risk and low-risk subjects (**Figure 3B**). Importantly, AIRE had similar performance in both men and women and in major ethnic groups (**Figure 3C** and **Table S2**).

### AIRE is superior to demographic data and traditional risk factors for mortality prediction

We compared the ability of **AIRE** to predict mortality, against using demographic data, risk factors and risk scores in the BIDMC test set (**Figure 3D**). **AIRE-Cox** had a significantly higher C-index than all other parameters combined (0.794 (0.792-0.795) vs 0.759 (0.758-0.761), p < 0.0001). Numerical results for all Cox models are shown in **Table S3**.

**AIRE-CV Death** predicted CV death with a C-index of 0.832 (0.831-0.834). In the BIDMC test set, **AIRE-CV Death-Cox** had a significantly higher C-index for prediction of CV death than all other parameters combined (0.844 (0.839-0.849) vs 0.795 (0.789-0.801), p < 0.0001) (**Figure 3E**). Finally, **AIRE-Non-CV Death** predicted non-CV death with a C-index of 0.749 (0.747-0.751). **Figure S2** shows sensitivity analyses using a single random ECG per subject.

### AIRE predicts mortality in transnational external datasets

First, we evaluated the performance of **AIRE** in the SaMi-Trop cohort of patients with chronic Chagas cardiomyopathy (11). Dataset demographics for all cohorts are shown in **Table 1**. The C-index was 0.773 (0.733-0.813, **Figure 4A**).

**Figure 4.**
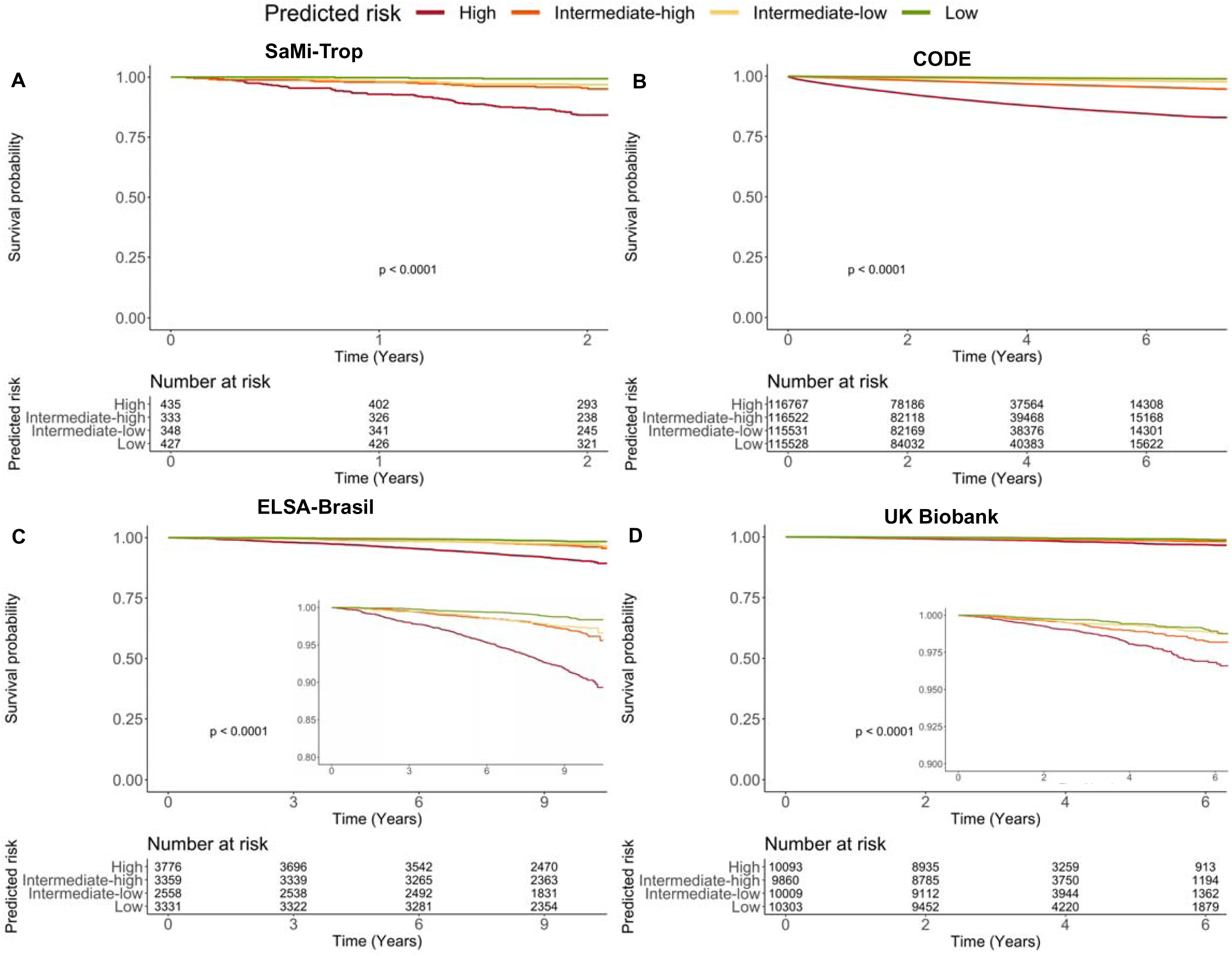
Survival analysis in four diverse, transnational, external validation datasets. In all cohorts, **AIRE/AIRE-primary care** successfully identified groups at higher risk of all-cause mortality. (A) São Paulo-Minas Gerais Tropical Medicine Research Center (SaMi-TROP) cohort of subjects with Chagas disease, **AIRE** predictions are shown, (B) CODE cohort of primary care subjects in Brazil, (C) The Brazilian Longitudinal Study of Adult Health (ELSA-Brasil) volunteer cohort, (D) UK Biobank volunteer cohort. AIRE-primary care results are shown for panels B-D. Sub panels show truncated y-axes for the populations with low event rates.

The CODE cohort is a Brazilian database of ECGs recorded in primary care (8). The dataset contains both 10s and 7s duration ECGs. We first evaluated the performance of **AIRE** without any fine-tuning. As **AIRE** was trained exclusively on 10s ECGs, we evaluated the model on the 10s subset (CODE-10s), **AIRE** had a C-index of 0.762 (0.759-0.765) for all-cause mortality prediction. **AIRE-primary care** more accurately predicted mortality with an improved C-index of 0.802 (95% CI 0.799-0.805, **Figure 4B**). When considering only the ECGs labelled as normal (26766 ECGs from 21897 subjects), there remained a significant difference in mortality between high-risk and low-risk subjects based on model predictions (**Table S1**).

Further evaluation of **AIRE-primary care** was performed in another independent external dataset, ELSA-Brasil (n = 13739) a volunteer cohort of civil servants from Brazil (12)). The C-index was 0.713 (0.691-0.735 **Figure 4C**). Again, when considering normal ECGs only there was a significant difference in mortality between high and low risk subjects (**Table S1**).

Finally, we additionally evaluated the performance of **AIRE-primary care** in the UK Biobank, a relatively healthy volunteer population (n = 42386) with only 526 (1.2%) deaths during follow-up. The C-index was 0.638 (0.608-0.668 (**Figure 4D**)) for all-cause mortality. As cause of death was available in the UKB, we additionally examined the ability of **AIRE-primary care** (which was trained for all-cause mortality) to predict CV death. C-index for CV death was 0.695 (0.636-0.754)

Hughes et al recently reported SEER (Stanford Estimator of ECG Risk) with the similar goal of predicting cardiovascular mortality (7). The model code and weights were downloaded, and performance evaluated in the UKB, as this was a dataset external to both **AIRE-primary care** and SEER, with cause of death available. **AIRE-primary care** was superior to SEER at predicting CV death in the UKB (SEER C-index 0.572 (0.514-0.630) p values for comparison to **AIRE-primary care** <0.001).

### AIRE outperforms AI-ECG models of biological aging

AI-ECG predicted age and the difference between AI-ECG predicted age and chronological age (“delta-age”) have been shown to be a better markers of biological aging than chronological age (8). We compared **AIRE/AIRE-primary care** predictions to AI-ECG predicted age. For the subsequent analyses, **AIRE** was used for SaMi-Trop and **AIRE-primary care** was used for CODE and ELSA-Brasil. We found **AIRE/AIRE-primary care**-predicted survival inversely correlated with AI-ECG age (CODE: r = −0.595, p < 0.0001, ELSA: r = −0. 449, p <0.0001, SaMi-Trop −0.479, p <0.0001) and, to a lesser extent, chronological age (CODE: r = −0.499, ELSA: −0.293, SaMi-Trop: −0.275, p < 0.0001 for all).

We compared the predictive capability of **AIRE/AIRE-primary care** with AI-ECG predicted age and delta-age in the CODE15 (a 15% subset of CODE), ELSA-Brasil and SaMi-Trop datasets. In age- and sex-adjusted models, **AIRE/AIRE-primary care** had a significantly higher C-index compared to AI-ECG predicted age or delta age in all three cohorts (CODE: 0.823 (0.813-0.833) vs 0.806 (0.796-0.816), ELSA-Brasil: 0.754 (0.734-0.774) vs 0.738 (0.718-0.758), SaMi-Trop 0.788 (0.749-0.827) vs 0.715 (0.672-0.758); p < 0.001 for all).

### AIRE performance in high-risk disease groups

For model predictions of mortality to be clinically useful, there needs to be specific interventions available to alter the trajectory of patients. Using the BIDMC test set, we investigated the performance of **AIRE** in two high-risk disease groups with existing risk stratification strategies and effective interventions. The data for high-risk disease group analysis was not available for the other cohorts.

For aortic stenosis, **AIRE** accurately predicted all-cause mortality, C-index: 0.701 (0.681-0.721, 1293 ECGs from 449 subjects). **AIRE-Cox** had a significantly higher C-index compared to all other parameters combined, **Figure 5A**, C-index 0.709 (0.688-7.30) vs 0.683 (0.661-0.705), p < 0.001).

**Figure 5.**
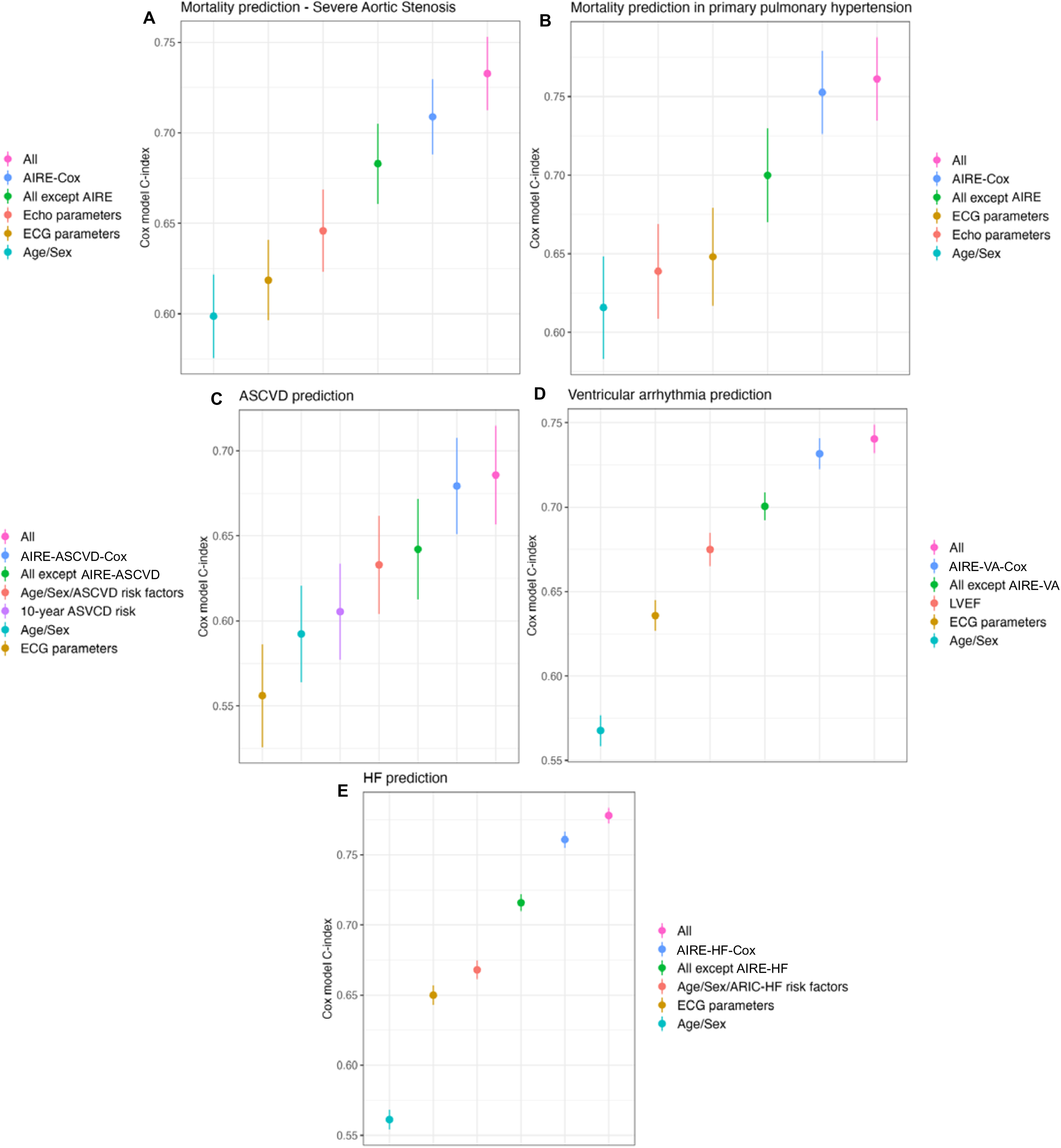
Mortality prediction in high-risk disease groups and prediction of actionable end-points - BIDMC Test set. Using Cox models, **AIRE** was compared with existing risk factors, ECG and imaging parameters in subgroups. In these Cox models, age, sex and ECG parameters were incorporated with **AIRE** to create **AIRE-Cox**. Severe aortic stenosis (A), and primary pulmonary hypertension (B). We also evaluated disease specific models, **AIRE-ASCVD** (atherosclerotic cardiovascular disease) (C), **AIRE-VA** (ventricular arrhythmia) (D) and **AIRE-HF** (heart failure) (E). Echocardiographic parameters for severe aortic stenosis: left ventricular ejection fraction (LVEF), aortic valve area, peak gradient and mean gradient. Echocardiographic parameters for primary pulmonary hypertension: LVEF, LV end diastolic diameter, tricuspid regurgitation (TR) pressure gradient, TR severity, right ventricular (RV) function, RV diameter. ECG parameters: heart rate, PR interval, QRS duration, QTc interval. ASCVD risk factors: systolic blood pressure, total cholesterol, HDL cholesterol, hypertension, smoking history, diabetes mellitus, ethnicity. 10-year ASCVD risk assessed using the pooled cohort equation. ARIC-HF risk factors: body mass index, systolic blood pressure, prevalent ASCVD, diabetes mellitus, smoking history, previous myocardial infarction, hypertension, ethnicity.

Similarly, for primary pulmonary hypertension, **AIRE** accurately predicted all-cause mortality, C-index 0.731 (0.724-0.738, 11741 ECGs from 789 subjects). In a subset with available recent echocardiograms (602 ECGs in 212 subjects, we evaluated **AIRE** in comparison and addition to other risk parameters. **AIRE-Cox** was superior all other parameters combined (C-index 0.753 (0.726-0.779) vs 0.700 (0.670-0.730), p < 0.005, **Figure 5B**).

### Actionable predictions: (1) ASCVD

**AIRE-ASCVD** was able to predict future ASCVD in subjects without known ASCVD (C-index 0.696 (0.694-0.698) n = 227588 ECGs from 56598 subjects). We externally validated these findings in the UKB Biobank (UKB); a healthy, volunteer population. **AIRE-ASCVD** had reduced performance in predicting ASCVD, C-index 0.643 (0.624-0.662) but was significantly better than the SEER model (7) (SEER C-index 0.547 (0.527-0.567), p <0.0001 for comparison with AIRE).

We compared **AIRE-ASCVD** to other risk parameters, including the pooled cohort equation (PCE) and ASCVD risk factors, in a subset of outpatients (4580 ECGs from 2926 subjects) in the BIDMC test set with appropriate available data. **AIRE-ASCVD-Cox** had a significantly higher C-index than all other factors combined (0.679 (0.651-0.708) vs 0.642 (0.613-0.672), **Figure 5C**, p < 0.00001). **Figure S3** shows sensitivity analyses using a single random ECG per subject for all three actionable prediction endpoints.

### Actionable predictions: (2) Ventricular arrhythmia

**AIRE-VA** was able to accurately predict future VA (C-index 0.760 (0.756-0.763) n = 393203 ECGs from 62443 subjects) in subjects without a previous history of VA. When compared to other conventional risk parameters, including ECG parameters and left ventricular ejection fraction, **AIRE-VA-Cox**, had a significantly higher C-index than all other factors combined (0.732 (0.723-0.741) vs 0.700 (0.692-0.709), **Figure 5D**, p < 0.0001). The **Supplementary Results** describe performance in subgroups of LVEF <50% and dilated cardiomyopathy.

In the UKB, (n = 34400 with both ECG and cardiac magnetic resonance imaging (CMR) and without previous VA, 44 events), **AIRE-VA** had similar performance in predicting first occurrence of VA 0.719 (0.635-0.803) and performed at least equivalent to LVEF from cardiac magnetic resonance imaging (CMR) (C-index 0.595 (0.494-0.697), p for comparison 0.11).

### Actionable predictions: (3) Future heart failure

**AIRE-HF** was able to accurately predict future HF in subjects without a previous history of HF (C-index 0.787 (0.785-0.889) n = 310200, from 61747 unique subjects). In a subset of patients with the available data, we compared **AIRE-HF-Cox** to other conventional risk parameters in Cox models (36486 ECGs from 12288 subjects), including HF risk factors identified in the Atherosclerotic Risk in Communities (ARIC) study (26). **AIRE-HF-Cox**, had a significantly higher C-index than all other factors combined (0.761 (0.755-0.767) vs 0.716 (0.710-0.722), **Figure 5E**, p < 0.00001). In the external validation cohort, UKB, **AIRE-HF-Cox** had similar performance at predicting future HF (C-index 0.768 (0.733-0.802).

### Single-lead ECG model performance

To evaluate if **AIRE** could be potentially used in wearable devices or inpatient telemetry units, we trained a single-lead version of **AIRE** using lead I only (**AIRE-1L)**. The performance of **AIRE-1L** (C-index 0.751 (0.750-752) **Figure S4**) was only slightly inferior in discrimination compared to the 8-lead **AIRE** model.

### Explainable ECG morphology associates with adverse prognosis

Using a variational autoencoder, we found features of QRS morphology, particularly broader and more left bundle branch block morphologies, inverted and biphasic T waves as well as ST segment changes were identified as the most significant morphological features associated with high predicted mortality (**Figure 6A**).

**Figure 6.**
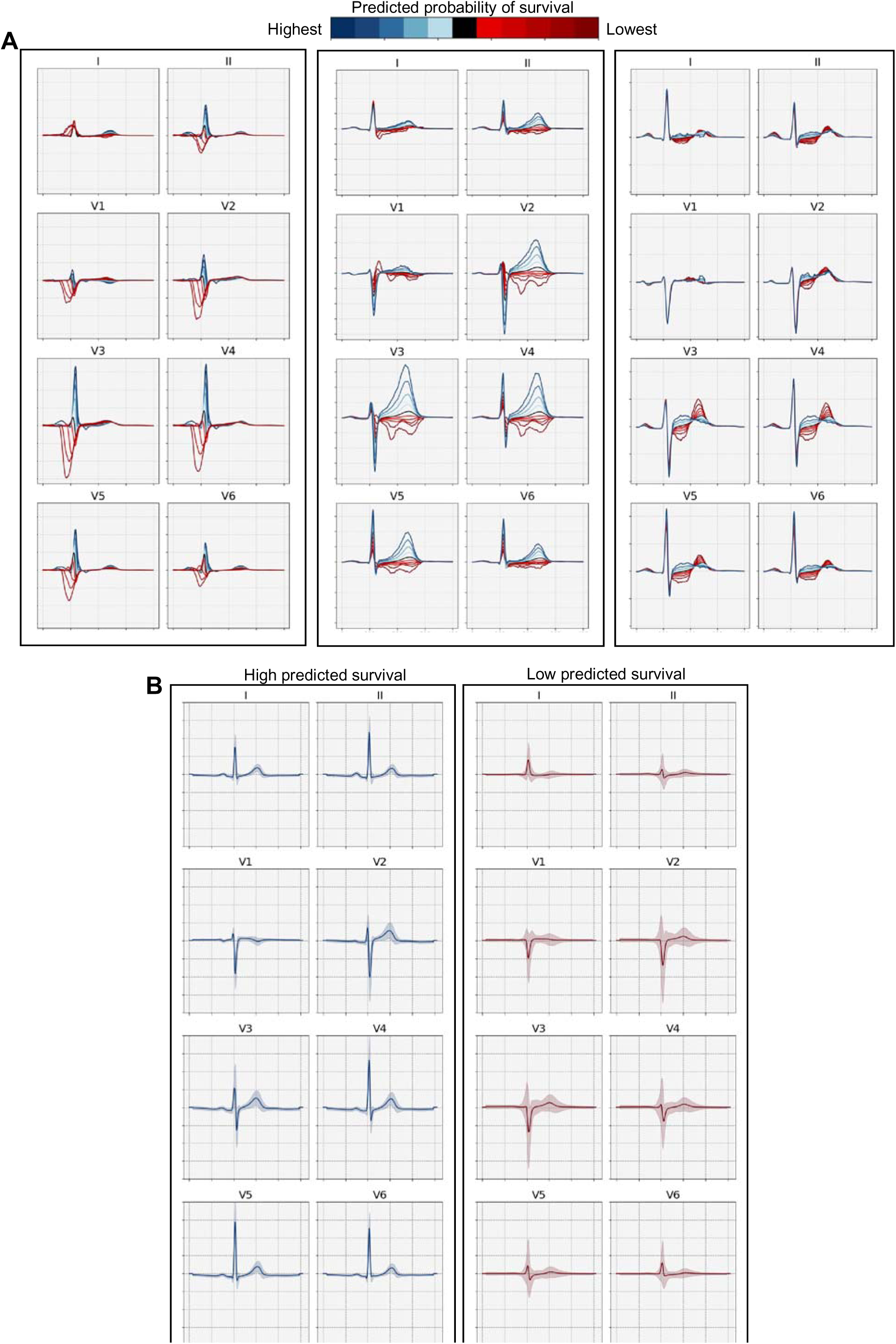
AIRE model explainability - BIDMC Test set: (A) A variational auto-encoder was used to identify the most important morphological features in **AIRE**-predicted mortality, each subpanel shows one of three latent features, identifying the importance of a broad QRS complex in a left bundle morphology as well as biphasic and inverted T waves. (B) Average ± standard deviation (shaded region) ECG waveforms for the 10,000 highest and lowest predicted survival ECGs from the BIDMC test set. This analysis identified poor R wave progression, low QRS amplitude and T wave flattening/inversion as important features in **AIRE**-predicted survival.

In a second approach, using median beats the BIDMC test set we found poor precordial R wave progression, low QRS amplitude and T wave flattening/inversion as important features in **AIRE**-predicted survival (**Figure 6B**).

### Biological plausibility: Genetic associations of AIRE-predicted survival

To investigate the underlying genetic associations with **AIRE**-predicted survival, we performed a genome-wide association study (**Figure 7A**). We found significant loci adjacent to *TBX3, VGLL2, CCDC91 and KCNQ1*. 8% of the total phenotypic variance in predicted survival was caused by the additive effects of genetic variation. *TBX3* has been associated with blood pressure (27), ECG morphology (28), myocardial mass (29) and trabecular development (24). *VGLL2* has been associated with ECG parameters, blood pressure, atrial fibrillation and BMI (27, 28, 30, 31). *KCNQ1* (Potassium Voltage-Gated Channel Subfamily Q Member 1) encodes K_v_7.1 and is most well-known for its associated with Long Qt Syndrome 1 and Jervell And Lange-Nielsen Syndrome 1 and QT interval (32) but is also associated with metabolic syndrome phenotypes (33–35). Finally, *CCDC91* (Coiled-Coil Domain Containing 91) associates with BMI (33), a variant associated with the ECG has been previously described in this region (28).

**Figure 7.**
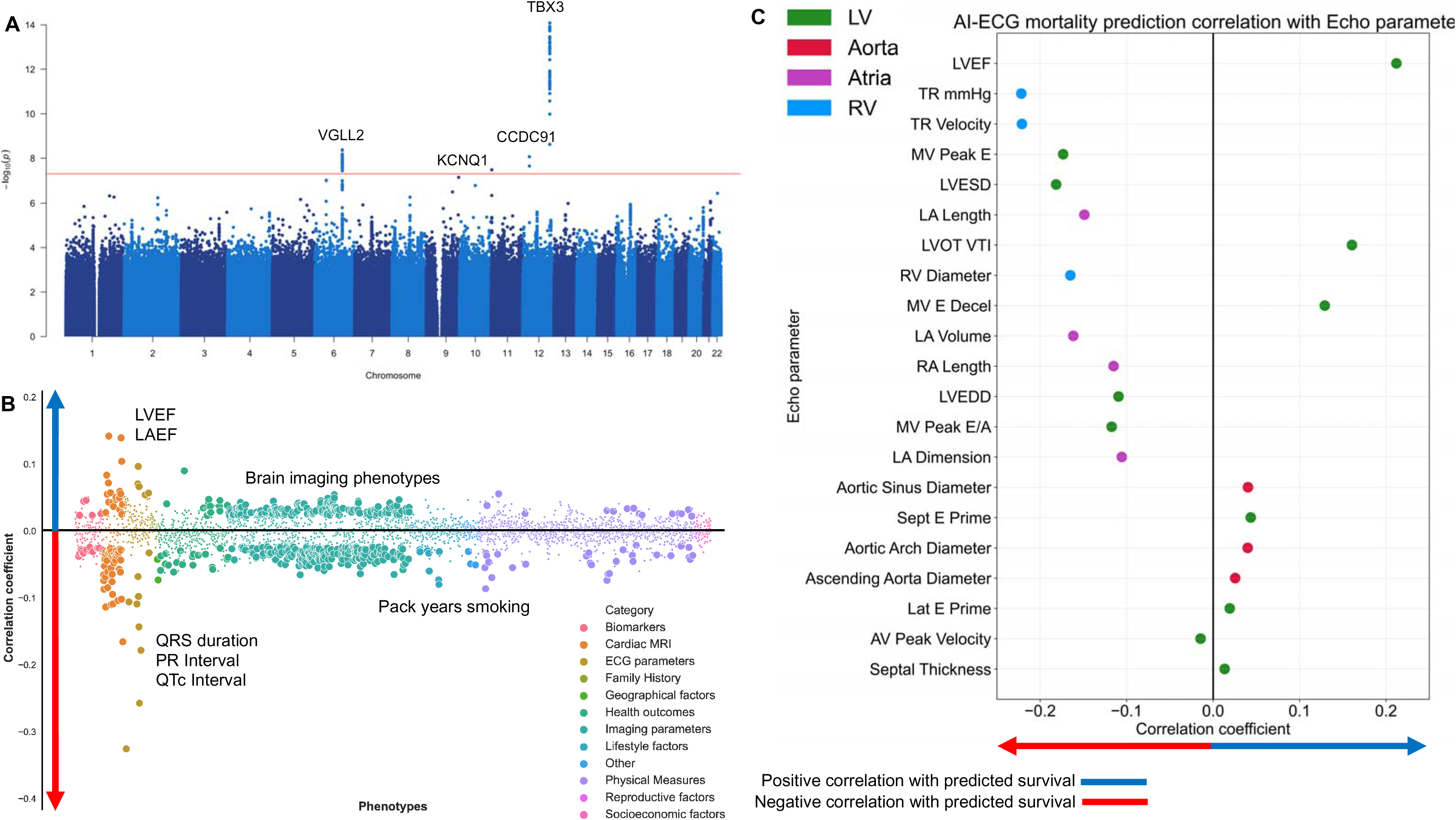
Exploration of underlying biology through Phenome and Genome-wide association studies. (A) Genome-wide association study (GWAS) in the UK Biobank. Manhattan plots of genomic loci associated with predicted survival. Nearest genes to significant single nucleotide polymorphisms are shown. The red line depicts the genome-wide significant threshold (P<5 x 10^-8^). (B) Phenome-wide association study in the UK Biobank. Cardiac associations include left ventricular ejection fraction (LVEF), atrial and right ventricular phenotypes. Non-cardiac associations included brain phenotypes such as total volume of white matter hyperintensities and pack years of smoking. (C) Association of **AIRE**-predicted survival with echocardiographic parameters in the BIDMC test set. LA: left atrium, LAEF: LA ejection fraction, TR: tricuspid regurgitation, MV: mitral valve, LVESD: LV end-systolic diameter, LVEDD: LV end-diastolic diameter, RA: right atrium, AV: aortic valve.

### Biological plausibility: Phenotypic associations of AIRE-predicted survival

In order to investigate the biological associations with **AIRE**-predicted mortality, we performed a PheWAS in the UK Biobank, **Figure 7B**. In particular, CMR associations included reduced left ventricular (LV) ejection fraction (EF), more positive, i.e., abnormal, global longitudinal strain, increased LV mass and increased left atrial size, which were correlated with reduced **AIRE**-predicted survival. We additionally specifically examined the association between predicted survival and left ventricular trabeculation and found a significant negative correlation (**Table S5**). Important lifestyle risk factors included smoking status and alcohol intake which were both correlated with reduced predicted survival. Significant physical measures included systolic blood pressure, which was negatively correlated with predicted survival.

In BIDMC echocardiographic analyses (**Figure 7C**), we found LVEF was positively correlated with predicted survival while LA volume and surrogate measures of pulmonary pressure (TR velocity) and right ventricular diameter were negatively correlated.

Finally, investigation of non-cardiac imaging phenotypes identified associations with multiple multi-model brain imaging phenotypes including the total volume of white matter hyperintensities and deep learning-derived brain age (25) (**Table S6**). Other non-cardiac imaging associations included predicted-survival inversely correlating with carotid intimal-media thickness, which is a marker of overall atherosclerotic burden (36).

## Discussion

We describe, for the first time, an actionable, explainable, and biologically plausible mortality and risk prediction AI-ECG platform of eight AI-ECG models. Importantly, our platform was externally validated across ethnically and demographically diverse transnational cohorts.

### Importance of time-to-event for actionability

This paper substantially extends the work of others on mortality protection using the ECG. Raghunath et al previously described the use of deep learning for mortality prediction (37), while Sun et al more recently built upon this work (6). Our study has several significant differences from these previous publications. Firstly, the use of a survival neural network architecture, provides our model with the ability to predict time of death without being constrained to a small number of time points.

Additionally, this allows us to use training data from subjects that were censored, for whom the time for death was not known. Specifically, we created a model that provides an individualised survival curve based on a single ECG. The ability to identify patients at risk of short-term mortality versus those at risk of long-term mortality is an important distinction. For example, patients at risk of short-term mortality may benefit from early escalation to intensive care settings, while those at risk of long-term mortality may need more detailed outpatient, follow-up or aggressive medical or interventional treatment.

Furthermore, by comparing our model to existing clinical risk factors and demographic information we have demonstrated the significant additive value of our model beyond traditional approaches. Finally, we performed external validation across diverse populations, demonstrating the wide applicability of our model platform.

### Actionable risk prediction in high-risk disease groups

By predicting risk of death in high-risk disease groups, we were able to provide potentially actionable information that could guide treatment decisions. We described how **AIRE** can be used to predict the risk of death in severe aortic stenosis and primary pulmonary hypertension. Severe aortic stenosis has a high mortality if left untreated. Current clinical guidelines in general advocate intervention based on echocardiographic measures and the patient’s symptoms (38, 39). However, there is increasing evidence that earlier intervention in asymptomatic individuals at high-risk of death may be of benefit, and there are randomised control trials currently investigating this approach (40, 41). Primary pulmonary hypertension also has a significant mortality rate (42). Medical therapy is available, and transplantation may be undertaken (43), therefore accurate risk stratification is paramount in guiding treatment selection for pulmonary hypertension patients. Mortality prediction using **AIRE** may be helpful in guiding treatments in both these conditions.

### Event-specific risk prediction

Using disease specific models, we showed the **AIRE platform** can also predict future cardiovascular events such as ASCVD, HF and VA, in addition to predicting mortality. ASCVD prediction is currently used extensively in international guidelines for decision-making around lipid lowering therapies. A 10-year ASCVD is calculated and can be used to consider initiation of primary prevention statin therapy (44). In order to assess ASCVD risk, various scoring systems are used, including the PCE, SCORE2 and QRISK3 (44–46). The ECG, despite being a cheap and readily available investigation, does not feature in any of these scoring systems. In this study, we have shown that **AIRE-ASCVD** can provide additional information that could improve ASCVD risk prediction and may be superior or at least additive to the existing PCE.

Our analyses are additive to those of Hughes et al (7), who recently describe SEER, an AI-ECG model for the prediction of CV death. **AIRE-primary care** and **AIRE-ASCVD** was superior to SEER for CV death and ASCVD prediction in the UKB. Additionally, the time-to-event nature of our platform provides additive actionable information as discussed above (7).

Similarly, predicting future VA is a particularly important endpoint, as there are clear preventative, and therapeutic options. For example, in-hospital predictions of short-term high risk of VA could be used to guide the need for cardiac monitoring and potentially even prophylactic use of antiarrhythmics. Medium-term risk could be managed with a wearable cardioverter defibrillator (47), while patients at longer-term risk of ventricular arrhythmias may benefit from primary prevention implantable cardioverter defibrillator implantation. Current guidelines advocate LVEF as the primary factor in determining eligibility for a primary prevention ICD. In our study, we demonstrated that **AIRE-VA** is a better predictor of future VT/VF than LVEF and could therefore potentially be incorporated into this decision-making paradigm.

Lastly, predicting future heart failure is important given the high number of unplanned hospital admissions due to undiagnosed heart failure (48). Early clinical assessment, echocardiography and institution of appropriate therapies may reduce adverse events, particularly in HF with reduced ejection fraction (HFrEF) (49).

### Single-lead ECG applications

Extending risk prediction to the single-lead ECG is particularly important given the rapidly increasing number of single lead devices, including consumer products (50). Others have shown the applicability of AI-ECG algorithms to consumer single-lead products (51). Our study highlights the excellent performance of **AIRE** at mortality prediction on only a single lead. This could be particularly applicable for inpatient cardiac monitoring, or for remote monitoring in the outpatient setting using wearable devices, for example in patients with chronic diseases such as heart failure, where high-risk predictions could trigger pre-emptive treatments to prevent hospital admissions.

### Explainability and biological plausibility

A major advantage of the deep learning approach is the ability to extract features relevant to the specific task, without anchoring on prior beliefs. However, a significant challenge is explainability of the model predictions. A significant barrier to the adoption of AI tools in clinical practice is physician reluctance to adopt technologies that are often “black boxes” (52). Explainability and biological plausibility is therefore key to improving clinician trust in AI. In this study using multiple approaches, we have explored the underlying biology behind **AIRE** risk predictions. Our ECG morphology findings are in line with prior studies that highlight these features as being prognostically importantly (53–55). Importantly, the significant predictive value of **AIRE**, even in normal ECGs, shows how deep learning can additionally make use of ECG morphological features that clinicians deem to be normal.

The accuracy of **AIRE** at mortality prediction raises the important question as to which biological pathways are being identified in these predictions. Using cardiac imaging data in two distinct datasets, we identified associations with cardiac chamber structure, function and measures of pulmonary pressure, that may be reflected in the ECG (56). We also identified multiple established factors, whose effects maybe reflected directly or indirectly in the ECG, including blood pressure, smoking and alcohol intake.

Previous work has described the importance of ischaemic heart disease on brain aging and outcomes (57). Our work has identified significant associations between ECG-predicted survival and brain imaging phenotypes that may reflect the combined effect of cardiovascular risk factors on cardiac and brain outcomes.

### AIRE-predicted survival is a biomarker of overall health

Through GWAS, we have identified plausible biological pathways, including myocardial mass and trabecular development, that could be a significant factors influencing **AIRE**-predicted survival. *TBX3* and *VGLL2* have been previously described in relation to AI-ECG derived delta-age, which is a marker of accelerated biological aging (8). Indeed, there was a moderate correlation between AIRE-predicted survival and AI-ECG predicted age, which was stronger than the correlation with chronological age. We also identified **AIRE**-predicted survival as inversely correlated with deep learning-derived brain-age. Additionally, we identified variants in *KCNQ1* and *CCDC91* that suggest **AIRE** may capture metabolic risk as an additional mechanism. These findings suggest **AIRE**-predicted survival is a biomarker of overall health, including biological age and the presence of clinical and subclinical disease.

### Reduced performance in volunteer populations

In our analyses, in general, **AIRE-primary care** had reduced performance metrics in the volunteer populations (UK Biobank, ELSA-Brasil). Our findings are consistent with other studies in that AI-ECG model performance is generally reduced in these volunteer populations (8, 58). A similar phenomenon is seen in other risk prediction models applied to the UK Biobank, such as QRISK3 (59). This may be due to differences in population characteristics, for example the UK Biobank population is older but healthier than the general population (60), and low event rates.

## Limitations

There are limitations to the accuracy and granularity of ICD diagnostic codes that are used in this study to ascertain disease status. In particular, ventricular arrhythmias as reported by ICD codes are not necessarily sustained or haemodynamically significant and therefore patients predicted to have these events would not necessarily benefit from an implantable cardioverter defibrillator. Further evaluation in an implantable cardioverter defibrillator cohort is needed. The GWAS findings should be considered hypothesis generating given the absence of a replication dataset. The GWAS and UKB PheWAS results were drawn from a population of predominantly European ancestry and may not apply to other populations. Despite these limitations, we have demonstrated that the **AIRE platform** is highly effective at predicting the timing of multiple outcomes in multiple, diverse, transnational datasets.

## Conclusion

In conclusion, we describe the **AIRE platform**, an actionable, explainable and biologically plausible AI-ECG risk estimation platform that has the potential for use worldwide across a wide range of clinical contexts, including primary and secondary care, for short- and long-term risk prediction at a population and disease-specific levels.

## Acknowledgments

This research has been conducted using the UK Biobank Resource under Application Numbers 48666, 40616 and 47602. The authors would also like to thank InSIGHT Core in the Center for Healthcare Delivery Science at Beth Israel Deaconess Medical Center for assistance in obtaining primary data.

## Data availability

SaMi-Trop cohort was made openly available (https://doi.org/10.5281/zenodo.4905618). The CODE-15% cohort was also made openly available (https://doi.org/10.5281/zenodo.4916206). Restrictions apply to additional clinical information on the CODE-15% and SaMi-Trop cohorts; to the full CODE cohort, the ELSA-Brasil cohort. UK Biobank data are available upon application (http://www.ukbiobank.ac.uk/). The BIDMC dataset is restricted due to ethical limitations. Researchers affiliated to educational, or research institutions may make requests to access the datasets. Requests should be made to the corresponding author of this paper. They will be forwarded to the relevant steering committee.

## Code availability

The programming code relating to these analyses will be made available under GNU General Public License version 3 upon request to the corresponding author.

